# Evidence Supporting EMA Drug Approvals (2020–2023): A Cross-Sectional Study of Trial Design and Outcomes

**DOI:** 10.64898/2026.02.04.26345500

**Authors:** Maximilian Siebert, Laura Caquelin, Florian Naudet, Joseph S. Ross, Reshma Ramachandran

**Author notes:** **Corresponding Author:** Maximilian Siebert, PharmD, PhD, Harvard-MIT Center for Regulatory Science, Harvard Medical School, Boston, Massachusetts, USA.

## Abstract

The strength and transparency of clinical trial evidence supporting drug approvals has become increasingly scrutinized, particularly considering the increased use of regulatory flexibility and expedited pathways. While U.S. Food and Drug Administration (FDA) standards have been extensively analyzed, evidence standards at the European Medicines Agency (EMA) remain less well-characterized. Thus, this study aims to systematically assess the design, quality, and outcomes of pivotal efficacy trials supporting EMA drug approvals between 2020 and 2023.

**Methods:** We conducted a cross-sectional analysis of new medicines and biosimilars receiving positive opinions from the EMA’s Committee for Medicinal Products for Human Use (CHMP) and subsequent approval by the European Commission between January 2020 and December 2023. Data were extracted from European Public Assessment Reports (EPARs) and EMA medicine databases. Key variables included trial design features, primary endpoint type and achievement status, and justification for approval in cases of failed efficacy endpoints.

**Results:** Between 2020 and 2023, 232 drugs were approved by the EMA for 281 indications. Of these, 205 (88.4%) were new active substances and 65 (28.0%) were granted orphan designation. Forty-six products (19.8%) were approved via a special regulatory program, most commonly Conditional Approval (26 products; 11.2%). Cancer was the leading therapeutic area, accounting for 61 approvals (26.3%). Approvals were supported by 393 pivotal clinical trials. Of these, 327 (83.2%) were randomized controlled trials (RCTs) and 218 (66.6% of RCTs) had a superiority design. A total of 232/393 trials (59.0%) relied on surrogate endpoints. Overall, 22 approvals (9.5%) were supported by at least one pivotal trial in which at least one primary endpoint was not met; in seven of these cases (31.8%), the failed trial was the sole pivotal trial. The most common rationale for approval despite null primary results was reliance on the totality of evidence, secondary endpoints, or clinical judgment (9 products; 40.9%).

**Conclusions:** Our findings reveal substantial variability in the design and evidentiary strength of pivotal trials supporting EMA approvals between 2020 and 2023. While the majority of studies were RCTs, reliance on surrogate endpoints was common. That 10% of approvals were based on pivotal trials with null primary endpoints highlights the nuanced role of regulatory judgment in therapeutic evaluation. These findings prompt reflection on evolving evidence standards in drug regulation and underscore the need for transparency and consistent justifications.

## Introduction

Balancing rapid access to innovative treatments against ensuring their safety and efficacy remains a central challenge for regulatory authorities when making drug approval determinations. Recent studies have highlighted the increasing use of flexible regulatory standards by the U.S. Food and Drug Administration (FDA), in part influenced by availability of special regulatory programs that expedite development and review, and utilize different evidentiary standards.^1–3^ For instance, one study found that drugs approved from 2015-2017 were more likely to use special regulatory programs and were based on fewer pivotal trials with less rigorous designs than drugs approved from 1995-1997.^2^ Another study found that between 2018 and 2021, 10% of approved indications were based on pivotal clinical trials that did not meet their primary efficacy endpoints, indicating potential compromises in the strength of evidence required for approval.^4^

Typically, the FDA emphasizes the necessity of at least two well-conducted randomized controlled trials (RCTs) to substantiate the efficacy of new drug applications ^5^. However, evidence suggests a growing flexibility in this standard. A recent study found that the proportion of indications supported by at least two pivotal trials declined from 80.6% in 1995–1997 to 52.8% in 2015–2017.^2^

These evolving practices raise important questions about how regulatory flexibility affects public health protection and how the FDA’s approach compares with international regulators. The European Medicines Agency (EMA), while facing similar pressures to expedite patient access, has often been viewed as applying stricter evidentiary standards. Yet comparative analyses suggest this distinction is narrowing. As such, between 1999 and 2014, 76 unique indications were approved without RCT results (44 by the EMA and 60 by the FDA), showing that both agencies have, at times, allowed substantial numbers of treatments to reach the market without traditional RCT evidence.^6^

While the FDA’s decision-making processes have been extensively studied, far less attention has been devoted to the EMA, leaving important gaps in understanding the evidentiary standards guiding its approvals. In particular, the frequency with which pivotal trials fail to achieve their primary endpoints in the European context remains unclear, and existing research has largely focused on specific therapeutic areas rather than providing a comprehensive overview. For instance, a 2019 study on cancer drugs approved by the European regulatory agency, found that nearly half of EMA-approved cancer drugs were at high risk of bias.^7^ Similarly, another analysis of psychotropic drugs approved by the EMA reported a lack of robust comparative effectiveness data: 81% showed no evidence of superiority over active comparators, only 8% demonstrated superiority in at least two such trials and clinical outcomes compared to placebo were generally modest.^8^ However, comprehensive analyses of a broader range of marketing approvals by the EMA remain limited.

Our objectives were to examine the design of clinical trials supporting EMA approvals, including the frequency of randomized controlled trials per approval, the categorization of drug types, the characterization of primary endpoints used in pivotal trials, and drawing on evidence that about 10% of FDA approvals involved failed primary endpoints, to quantify how often EMA-approved drugs were supported by pivotal studies that did not achieve their primary endpoints.

## Methods

### Study design

We used a cross-sectional study design to analyze new medicines and biosimilars that received positive opinions from the EMA’s Committee for Human Medicinal Products (CHMP) between 2020 and 2023 and were subsequently approved by the European Commission (EC).

### Data sources and identification of medicines

One researcher (MS) systematically reviewed CHMP agendas and minutes from 2020 to 2023 to identify eligible new drugs and biosimilars. Medicines with positive CHMP opinions were initially identified through the EMA’s official website. Subsequently, European Public Assessment Reports (EPARs) were manually retrieved corresponding from the EMA’s search portal.

EPARs are published on the EMA website upon final adoption by the EC and contain a detailed section on Main Studies, which are equivalent to pivotal trials in FDA approvals. For consistency with FDA terminology, we refer to these studies as pivotal trials from here on.

### Eligibility criteria

Medicines were included if (i) they received a positive EMA approval between 2020 and 2023, (ii) were subsequently approved from the EC post-positive CHMP opinion, (iii) were categorized as new drugs or biosimilars and (iv) contained at least one pivotal trial. We included biosimilars because their regulatory approval also depends on pivotal comparative trials. However, since these trials typically aim to establish equivalence rather than de novo efficacy, their evidentiary role differs from that of trials supporting new drugs. Medicines were excluded from the analysis for withdrawn or refused applications and for those categorized as generic or hybrid medicines.

### Data extraction

Two researchers (LC & MS) extracted data from the EMA website and the respective EPARs. From each EPARs, information was collected on: the number of pivotal trials studies, total number of primary endpoints, number of randomized controlled trials, blinding status, randomization and presence of control groups across all pivotal trials, number of patients enrolled, study design (superiority, non-inferiority, equivalence, or not applicable), outcome types (clinical, surrogate or clinical scale), primary endpoint achievement status. For drugs with at least one pivotal trial that did not meet one of its primary endpoints, we extracted and categorized the justification for marketing authorisation despite the missed endpoint.

Additional data were cross-checked using the EMA’s medicine data download webpage.^9^ These included the year and month of EMA opinion, drug name and International Nonproprietary Name (INN), orphan designation status, expedited approval status including special regulatory programs such as Priority Medicines (PRIME), Conditional Approval, Accelerated Assessment or Exceptional Circumstances (see Box for details), the pharmaceutical company or applicant, drug type (new medicine or biosimilar), therapeutic area (e.g., cancer, neurology) and specific indication (e.g., non-small cell lung cancer). Furthermore, we captured whether the approved products were advanced therapies. (i.e. medicine for human use that is based on genes, cells or tissue engineering) and whether it received additional monitoring (aims to enhance reporting of suspected adverse drug reactions for medicines for which the clinical evidence base is less well developed).

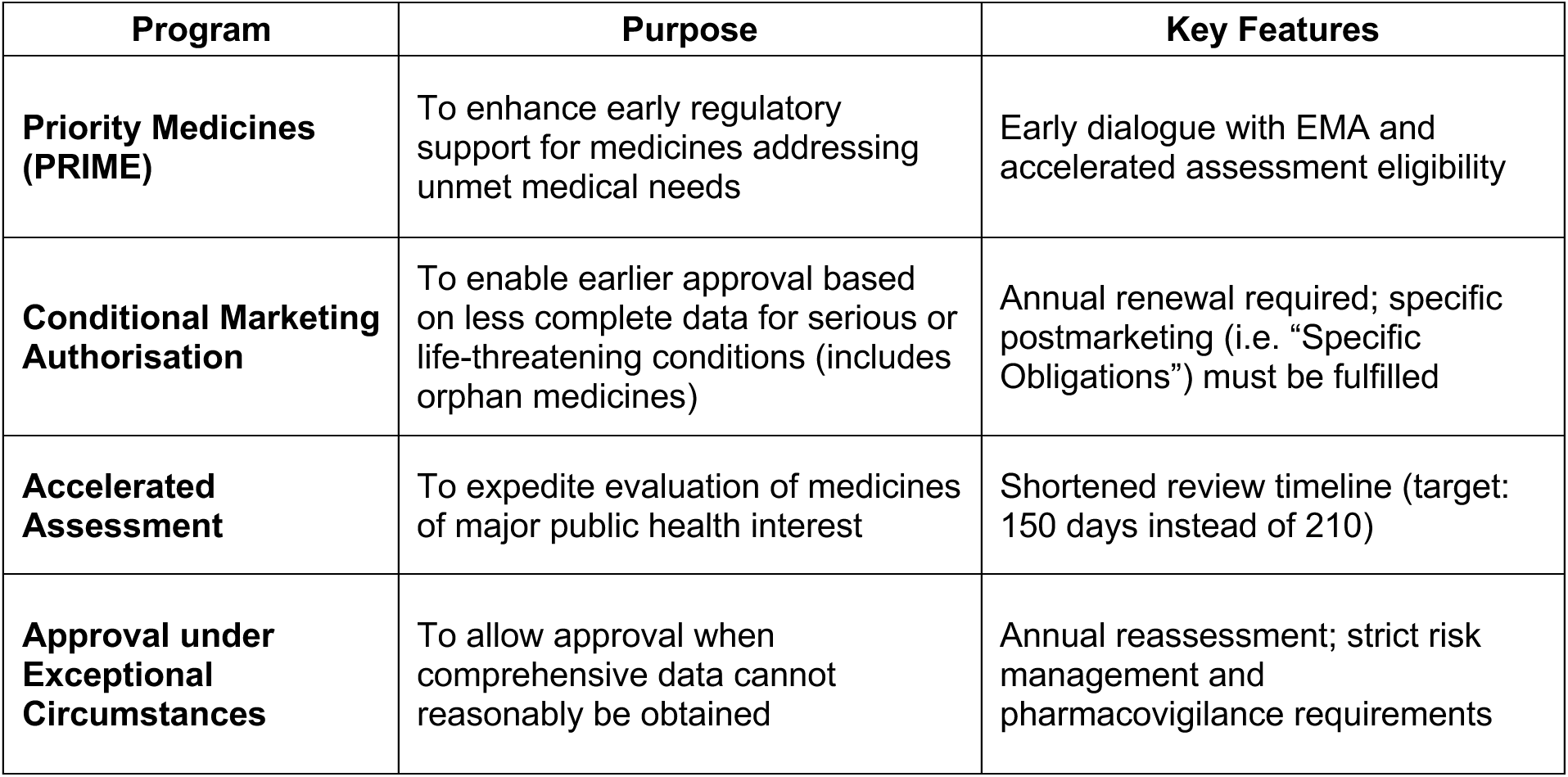

Box: Explanation of EMA’s special regulatory programs

### Analysis

Data were analyzed descriptively, presenting counts, percentages, medians and interquartile ranges. Qualitative analysis included verbatim quotes related to primary outcomes. All statistical analyses were conducted using R software version 4.0.5.

## Results

### Main Results

We identified 261 new drugs and biologics approved by the EMA. After exclusion of 21 drugs that were withdrawn or revoked by the time of our analysis and 8 drugs that did not include any pivotal trial, our sample contained 232 therapeutic products.

These 232 drugs accounted for 281 approved indications, as 14 drugs were approved for multiple indications: 4 drugs had two indications, 6 had four indications, 1 had five, 1 had 8 and 2 had nine indications.

Of the 232 drugs, 205 (88.4%) were new active substances compared to 27 (11.6%) biosimilars (Table 1). The EMA had granted orphan status to 65/232 (28.0%) products, and 46/232 (19.8%) were approved through any special approval pathway with Conditional Approval being the most used (26/232, 11.2%).

**Table 1.** Characteristics of 232 Drugs Approved by the European Medicines Agency (EMA), 2020-2023.

| **Drug characteristic** | **Number of Drug (%)**<br>**[N = 232]** | **Number without failed primary endpoints (%)**<br>**[N = 210]** | **Number of drugs with at least one failed primary endpoint (%)**<br>**[N = 22]** |
| --- | --- | --- | --- |
| Drug type |  |  |  |
| New active substances | 205 (88.4) | 186 (88.6) | 19 (86.4) |
| Biosimilar | 27 (11.6) | 24 (11.4) | 3 (13.6) |
| Special Regulatory Program |  |  |  |
| Any | 46 (19.8) | 41 (19.5) | 5 (22.7) |
| Accelerated Assessment | 3 (1.3) | 2 (1.0) | 1 (4.5) |
| Conditional Approval | 26 (11.2) | 25 (11.9) | 1 (4.5) |
| Exceptional Circumstances | 11 (4.7) | 8 (3.8) | 3 (13.6) |
| PRiority Medicines (PRIME) | 21 (9.1) | 20 (9.5) | 1(4.5) |
| None | 186 (80.2) | 169 (80.5) | 17 (77.3) |
| Orphan drug | 65 (28.0) | 63 (30.0) | 2 (9.1) |
| Advanced therapy | 10 (4.3) | 9 (4.3) | 1 (4.5) |
| Additional monitoring | 188 (81.0) | 170 (81.0) | 18 (81.8) |

Out of the 232 therapeutics, 10 (4.3%) were advanced therapies and the majority, 199/232 (81.0%), received the label of additional monitoring.

The most prevalent therapeutic area was Cancer with 61/232 (26.3%) agents, followed by Neurology and Haematology/Haemostaseology, with 27/232 and 18/232 drugs respectively (11.6% and 7.8%) (Figure 1).

**Figure 1.**
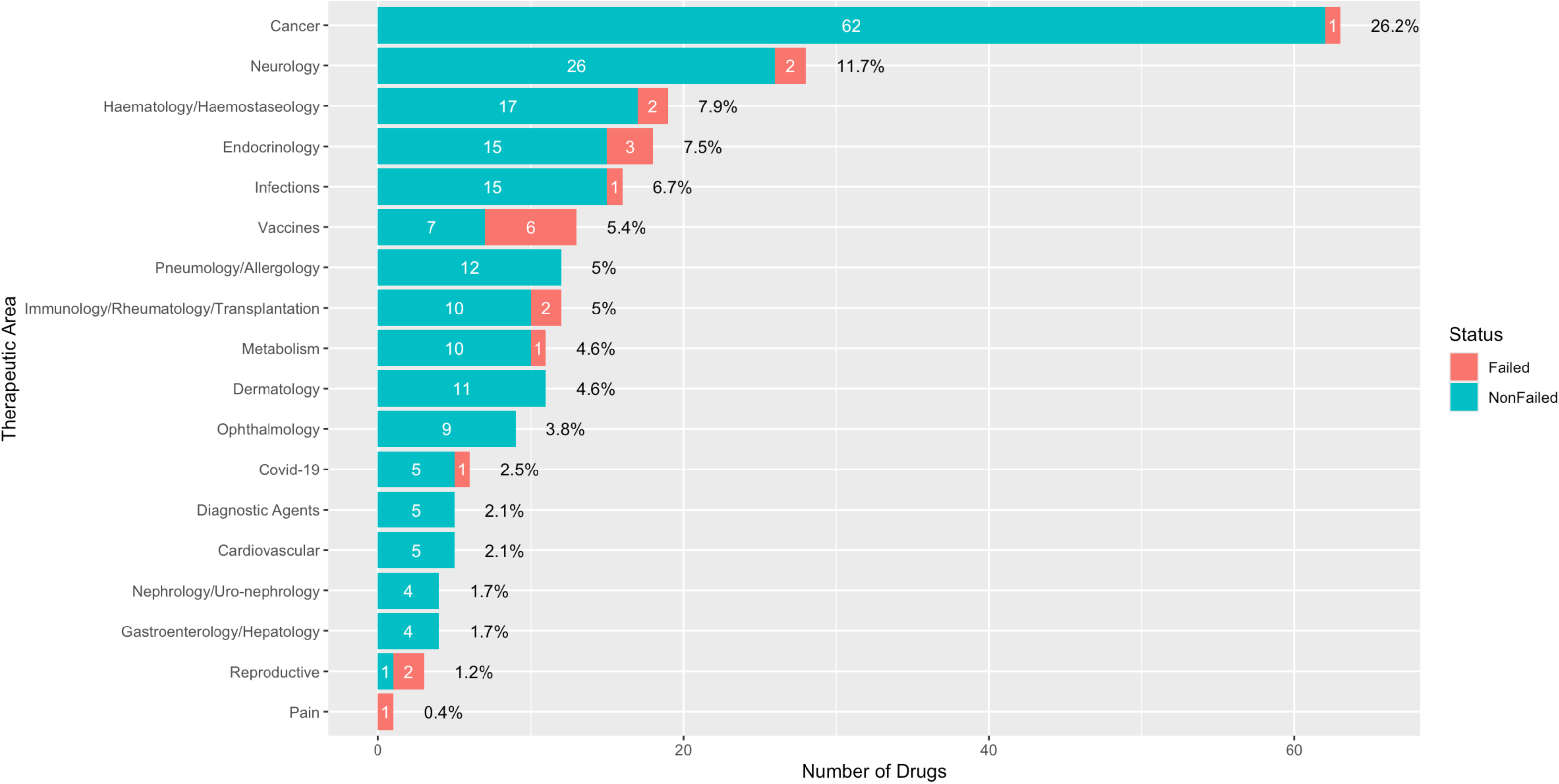
Distribution of 232 Drugs approved by the European Medicines Agency (EMA) by Therapeutic Area and Primary Endpoint Outcomes, 2020-2023 The horizontal bars represent the total number of drugs approved within each therapeutic area. The blue segments indicate drugs not supported by pivotal trials with null findings for any primary endpoints. The red segments represent drugs supported by pivotal trials with null findings for one or more primary endpoints. Percentages at the end of each bar indicate the proportion of the total drug count represented by each therapeutic area.

### Main Efficacy Study Features

These 281 approved indications were based on a total of 393 pivotal trials. The median number of pivotal trials per approval was 1.0 (interquartile range [IQR], 1.0-2.0) and the median number of patients enrolled per study was 449 (IQR, 162–913). 60.3% (140/232) based on one single pivotal trial.

The primary endpoint was a surrogate outcome for 232/393 trials (59.0%), a clinical outcome for 115/393 (29.3%) trials, and a clinical scale for 42 (10.7%) trials. For four trials, a combined endpoint was found, where one contained a mix between a clinical scale and surrogate endpoint (0.2%), and three combined a clinical and surrogate endpoint (0.8%).

The majority of pivotal trials, 327/393 (83.2%), were RCTs and 254/393 were double-blinded (64.6%). Out of 327 RCTs, 218 (66.6%) trials had a superiority design, 59 (18%) non-inferiority and 28 (8.6%) equivalence design and 14 trials (4.3%) had no design or were exploratory. Additionally, 8 (2.5%) trials reported mixed designs: combining superiority and non-inferiority (6), or non-inferiority and superiority (1), or equivalence and non-inferiority (1).

### Drugs and trials that have not met their endpoints

Out of 232 drugs supported by at least one human clinical trial, 22 (9.5%) had at least one pivotal trial in which at least one primary endpoint was not met, corresponding to a total of 31 pivotal trials. The majority of these trials (64.5%; 20/31) assessed only one primary endpoint. For 7 of the 22 drugs (31.8%), the failed study was the only pivotal trial supporting the respective approval. In all 7 cases, the study contained only one primary endpoint, which was not met (see Table 2).

**Table 2.**
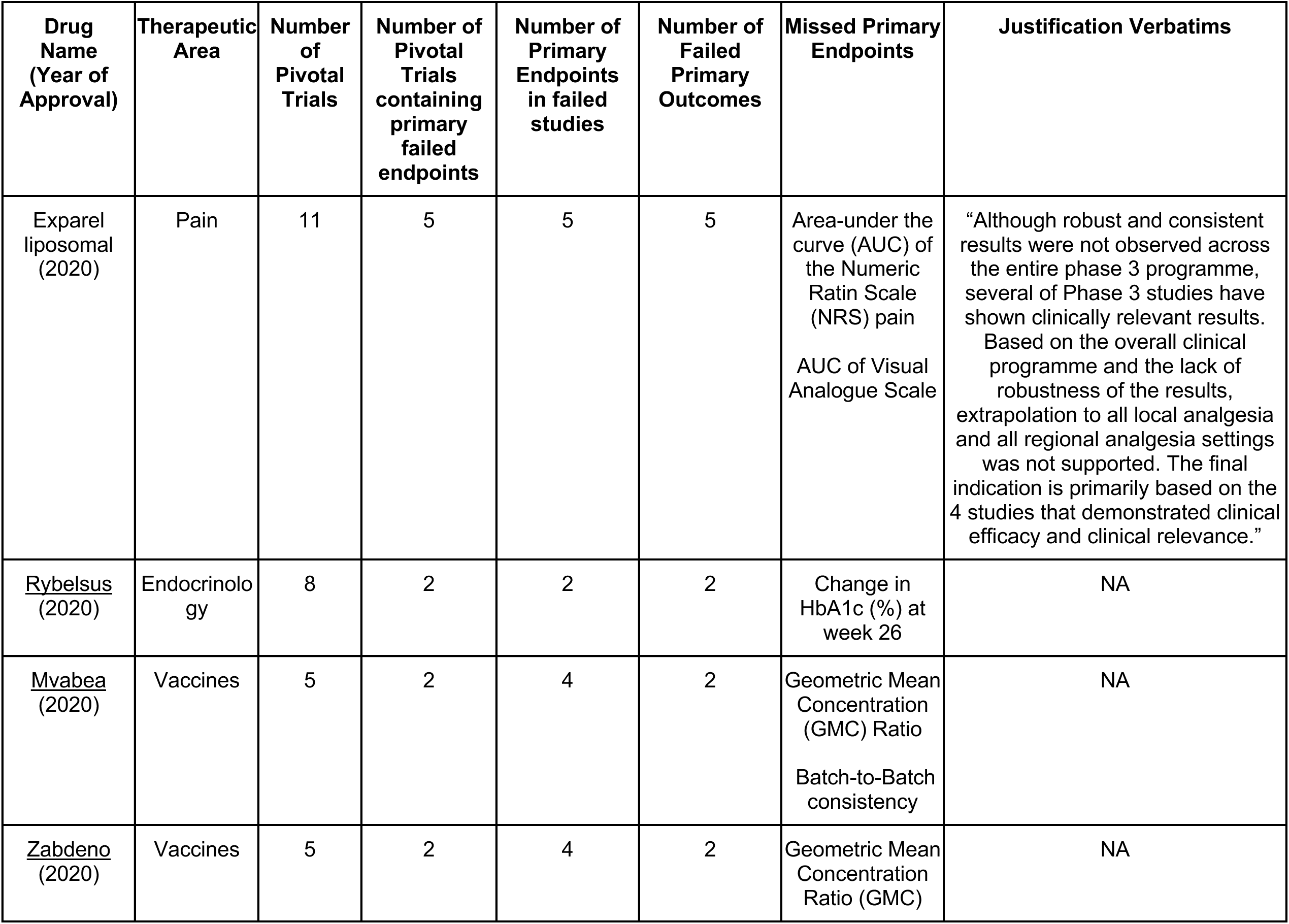
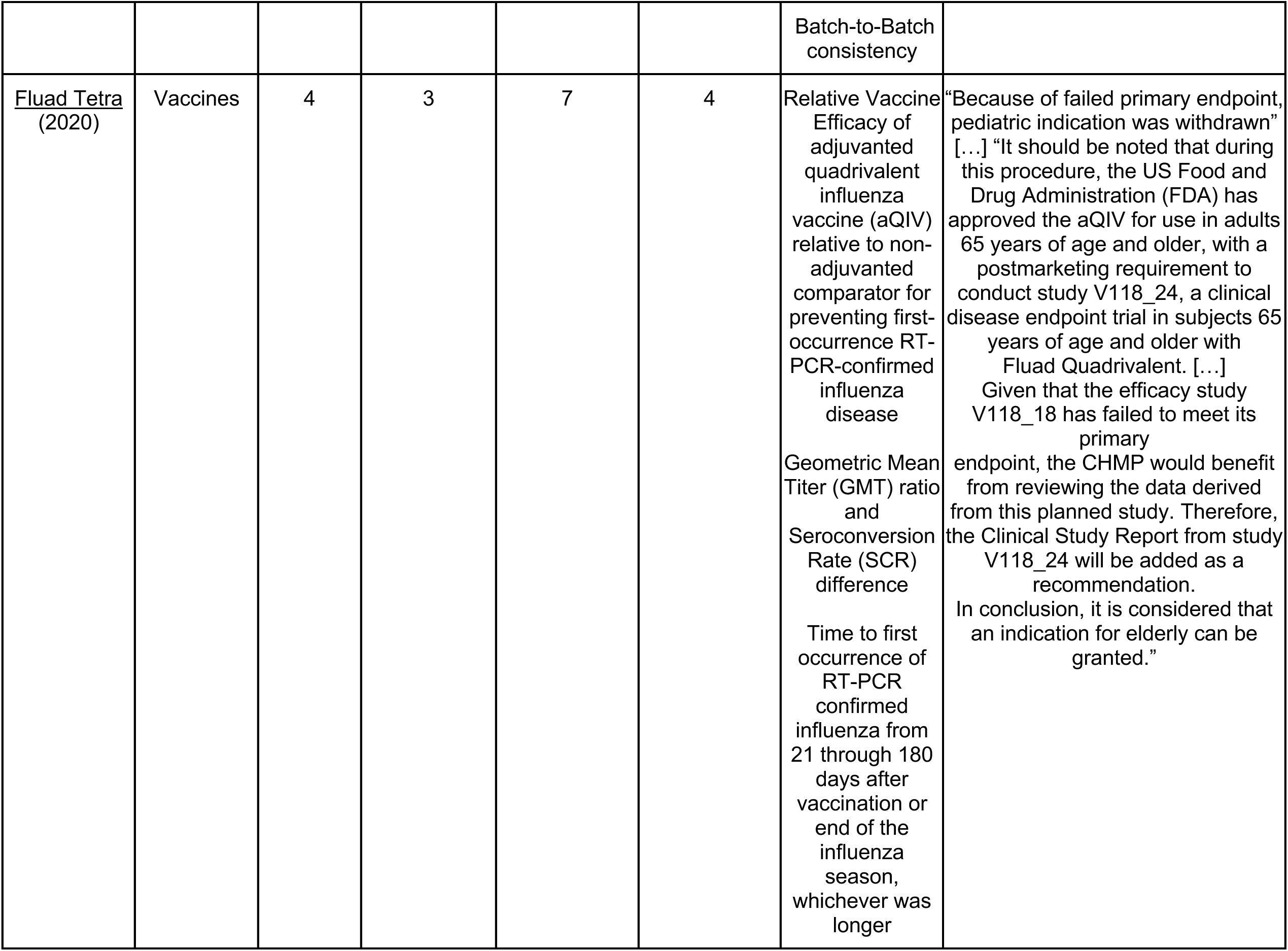
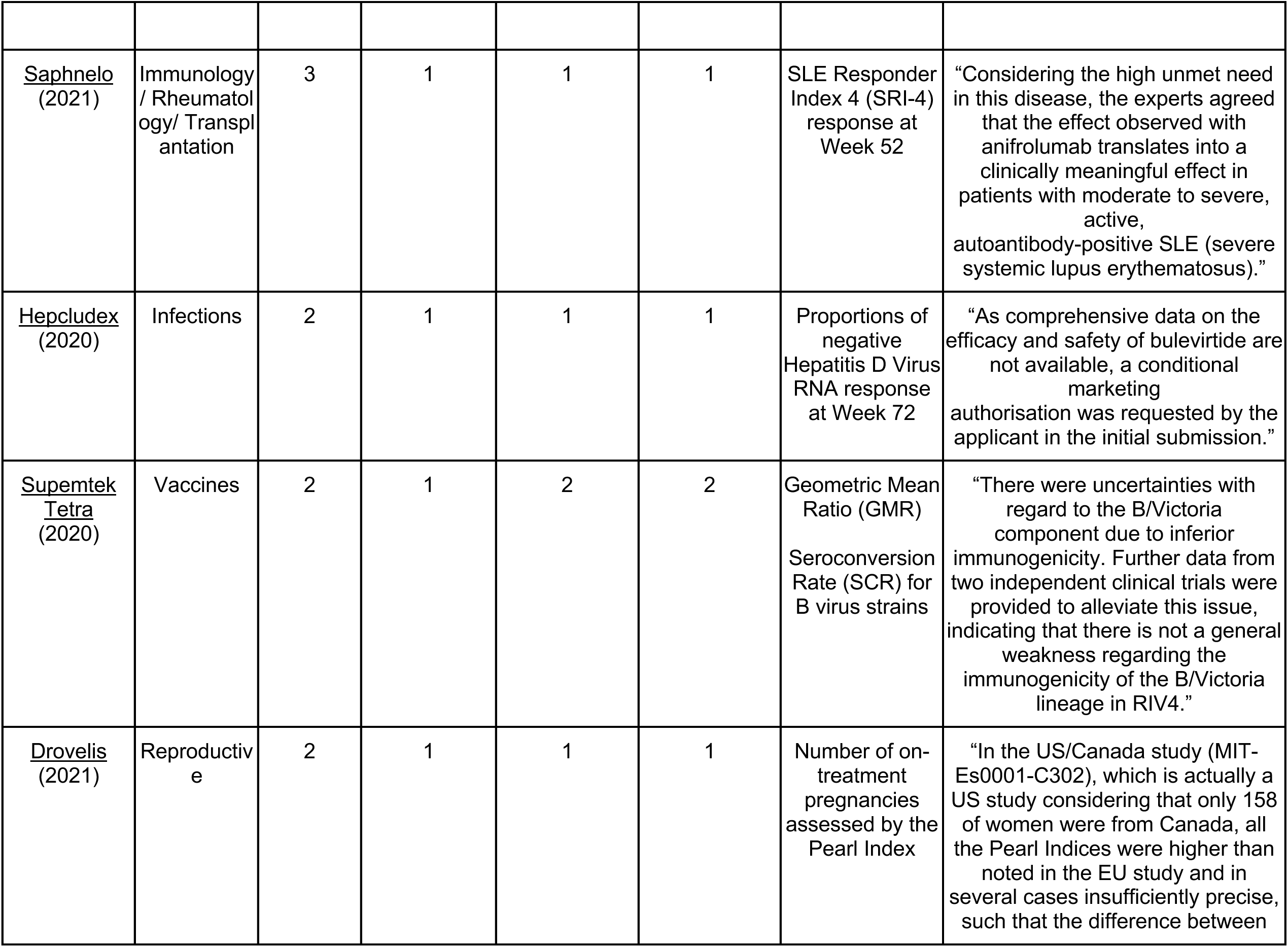
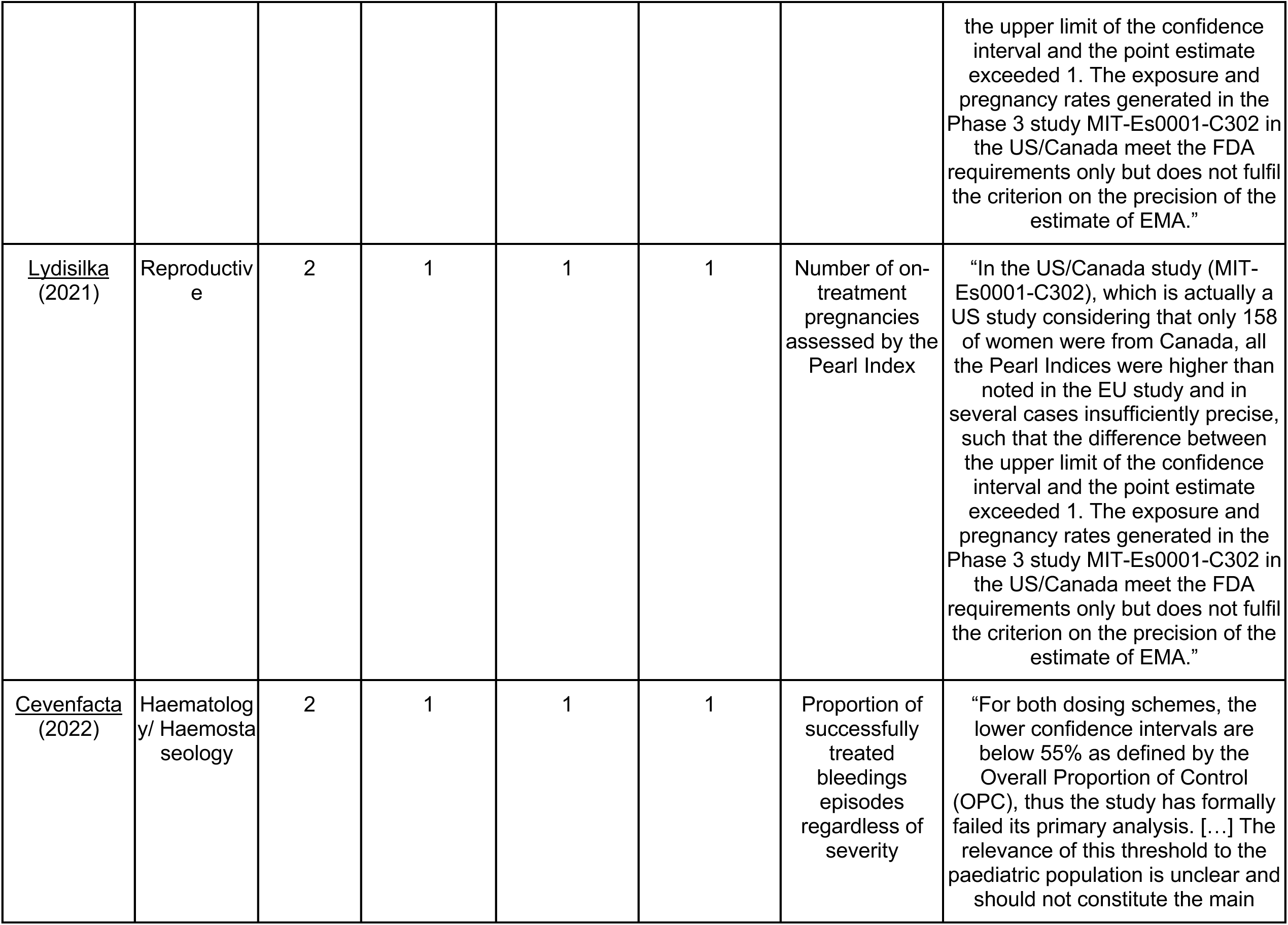
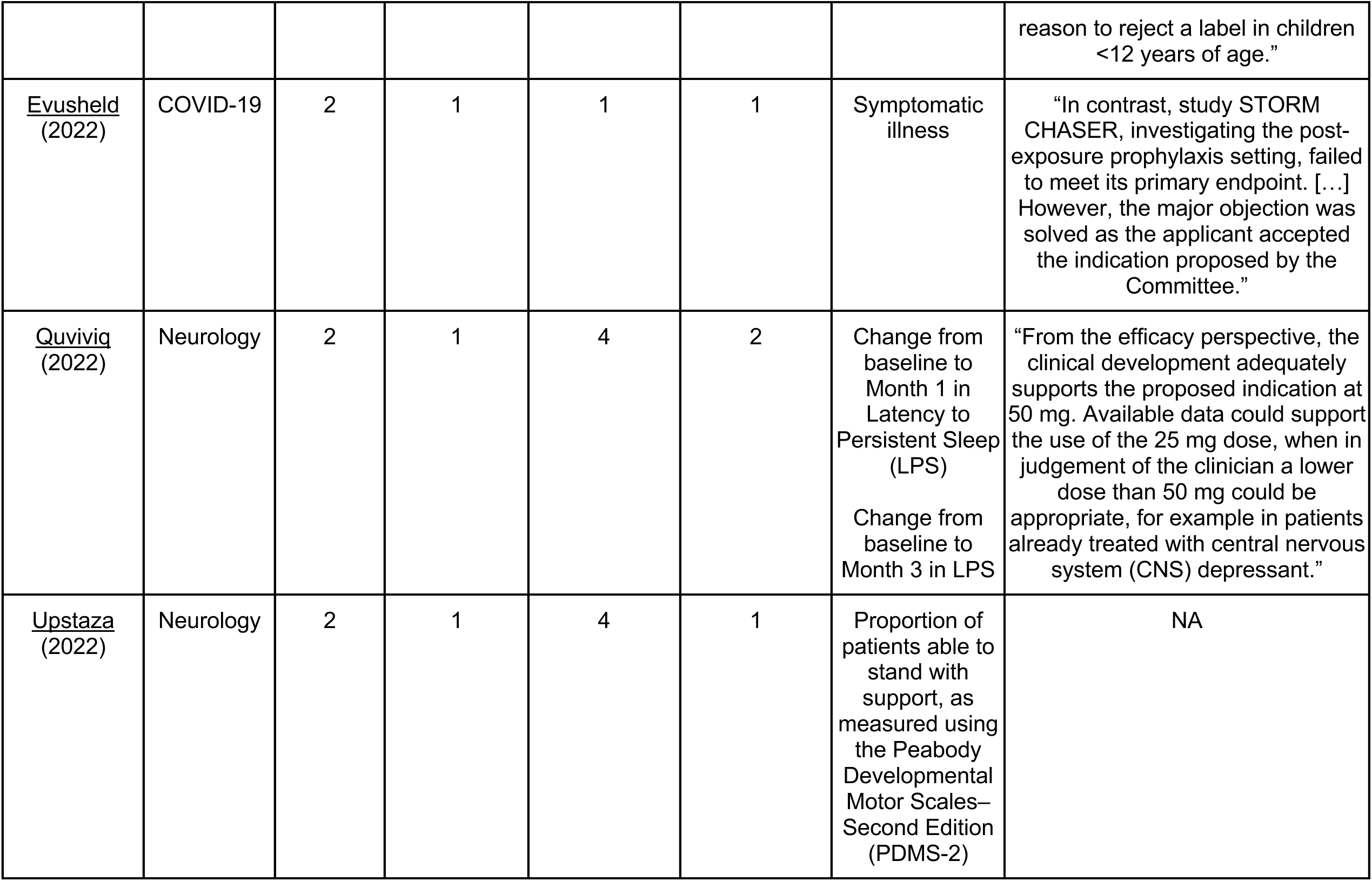
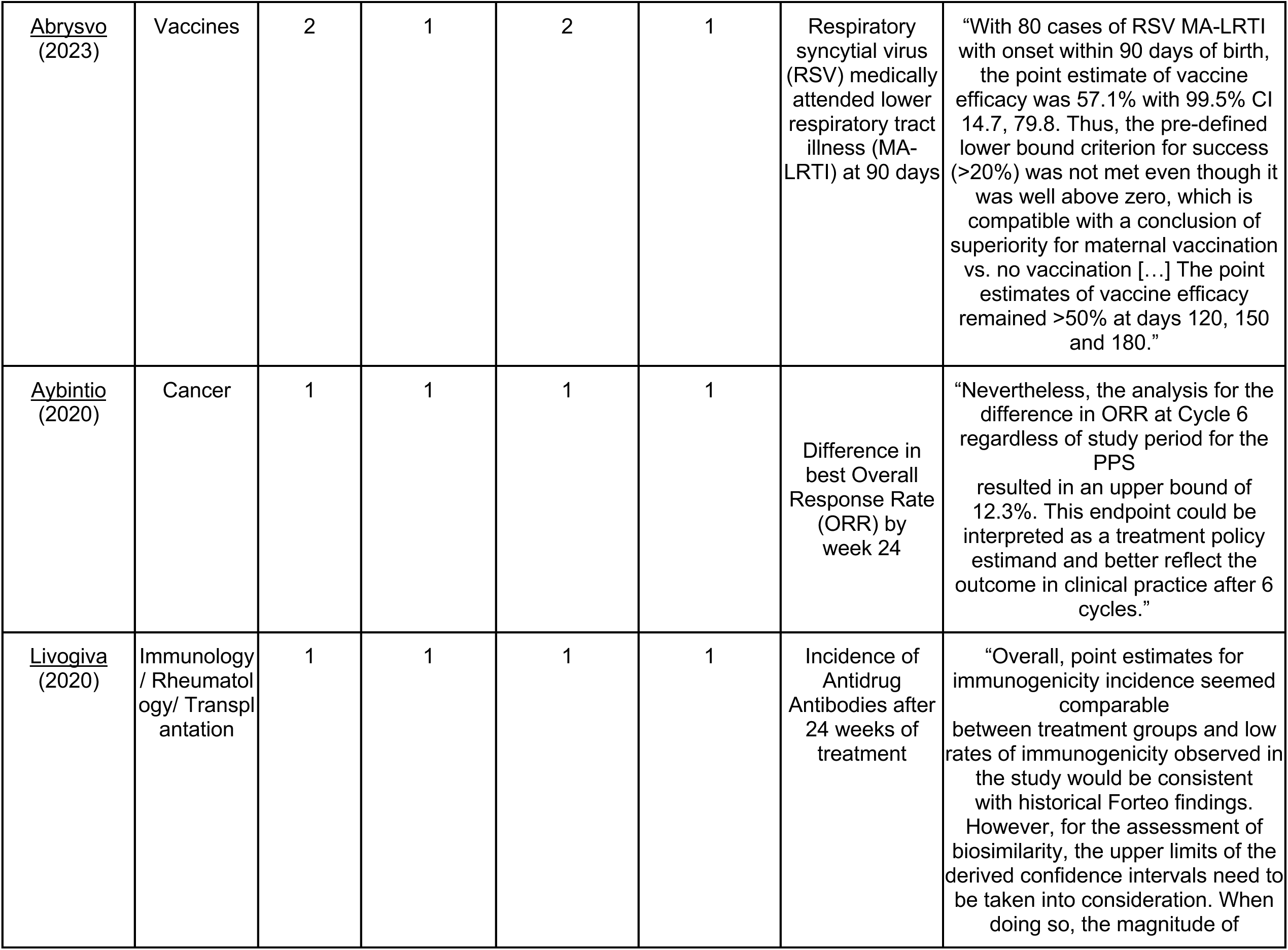
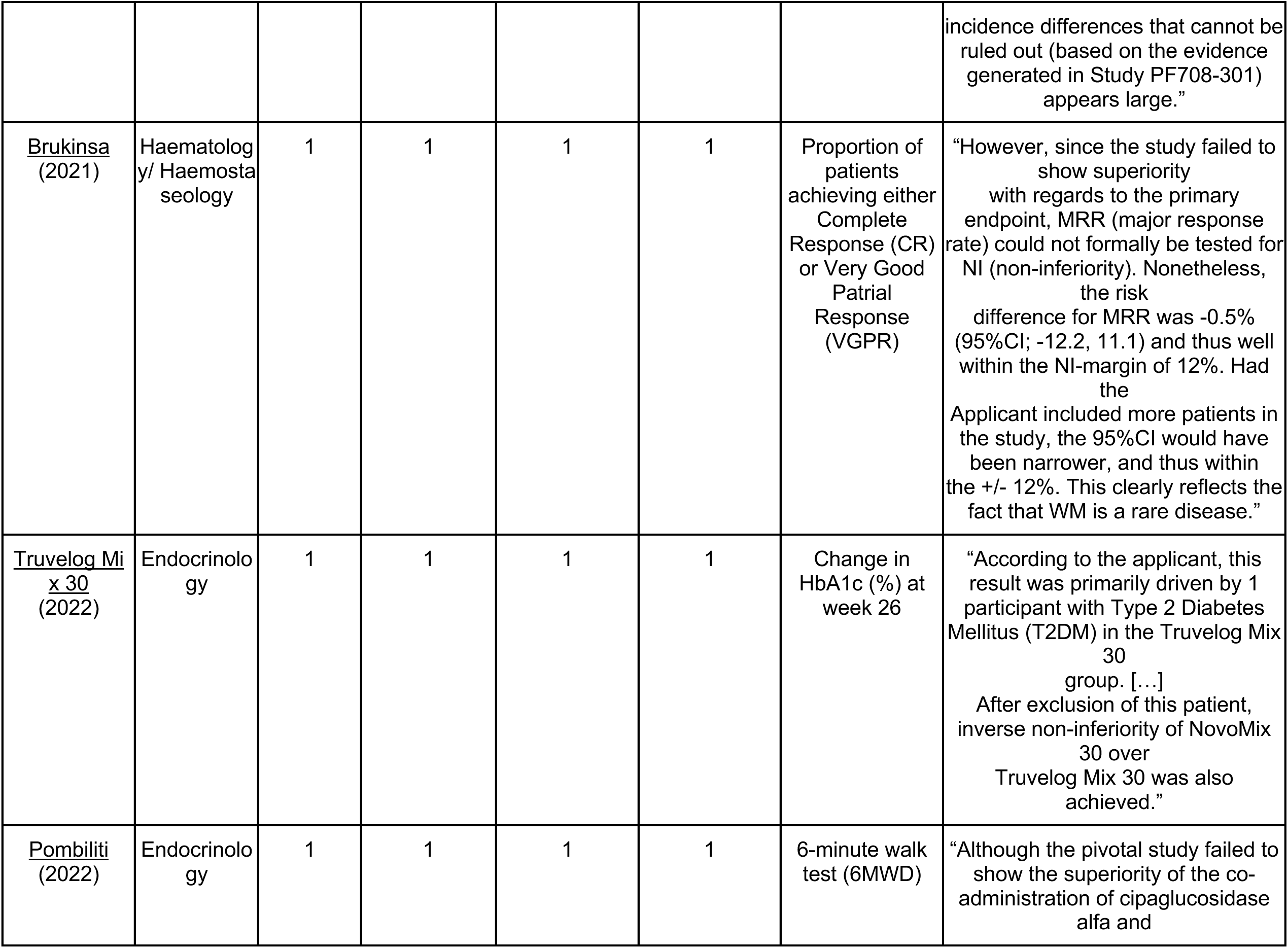
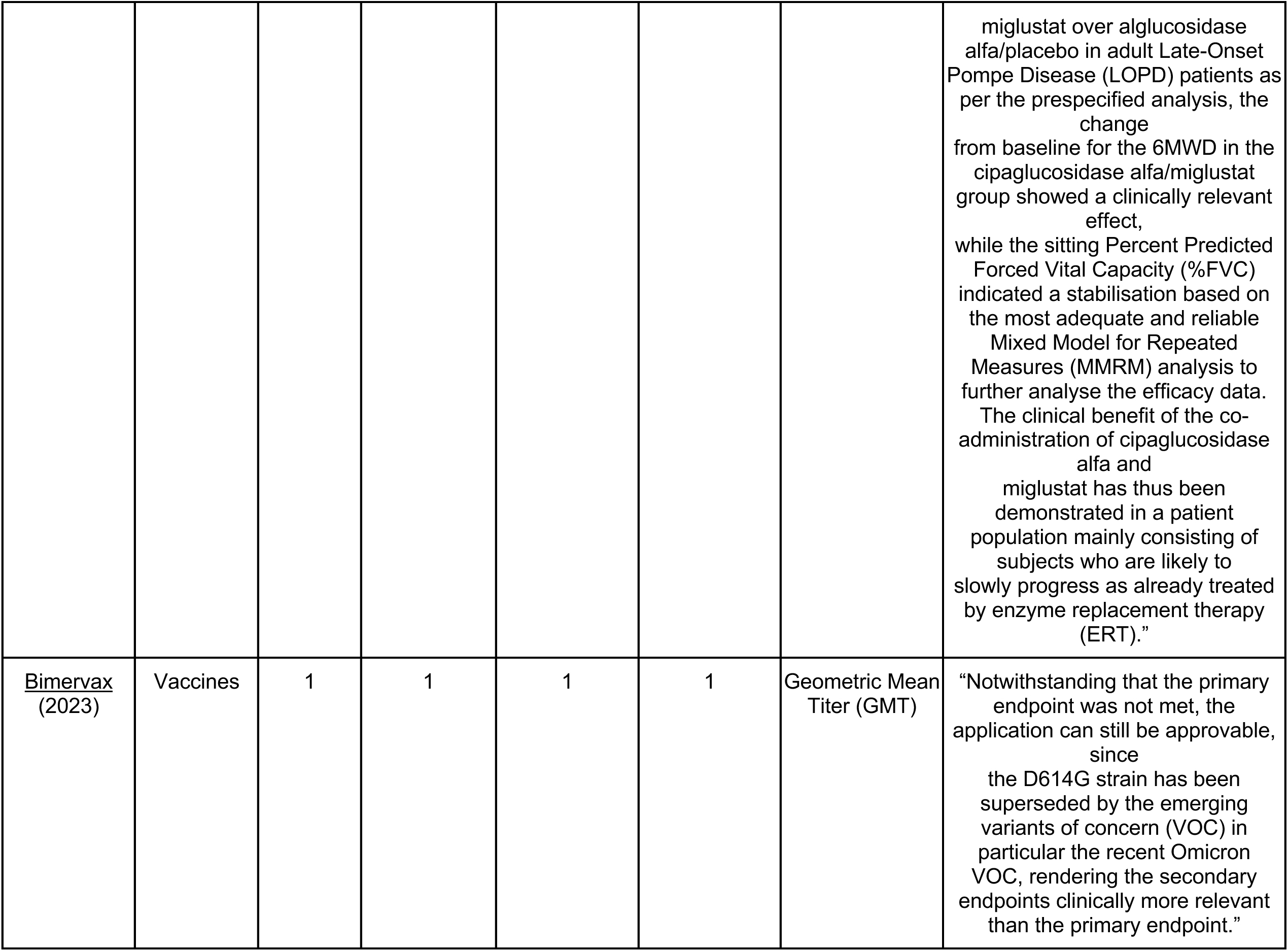
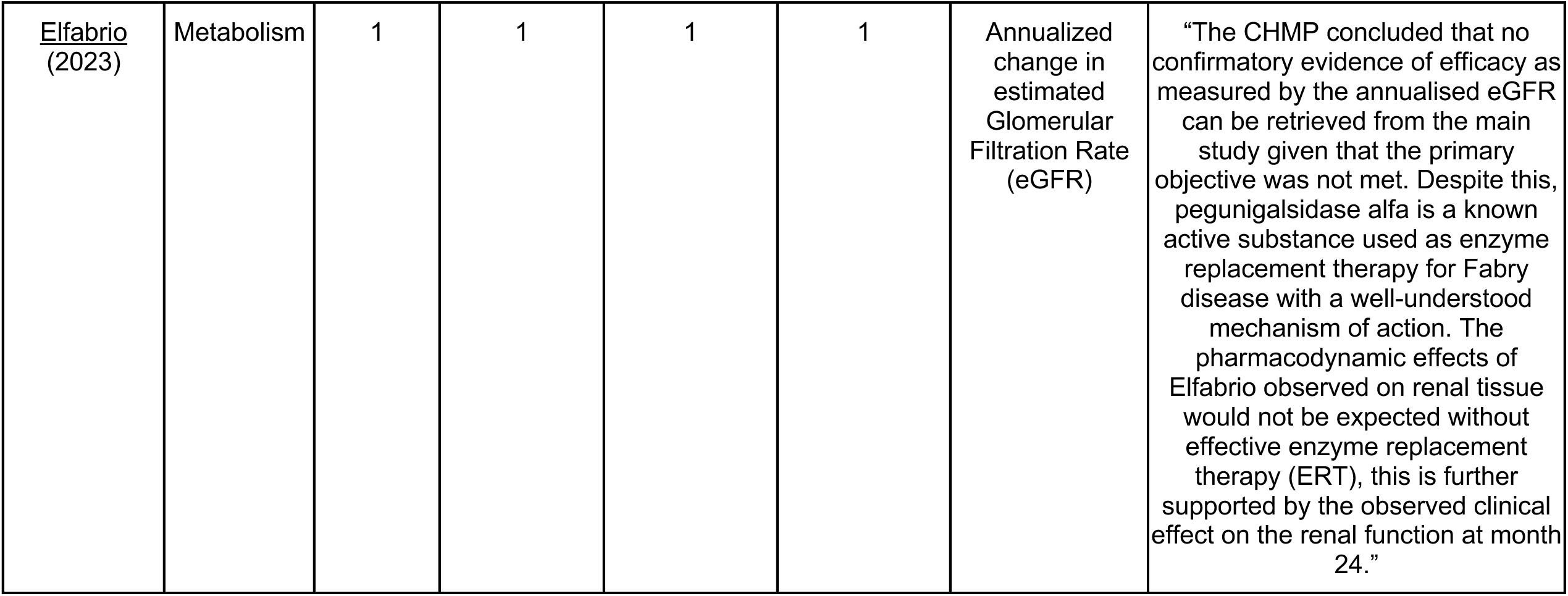
Descriptions of the 22 Drugs Approved by the European Medicines Agency (EMA) Supported by Pivotal Trials With Null Findings for 1 or More Primary Endpoints, 2020-2023.

Additionally, five drugs contained 2 or more primary endpoints. The most common therapeutic area was vaccines, which comprised 6 out of the 22 drugs (27.2%). For the majority, these were under additional monitoring at the time of approval (81.8%).

Among the 22 drugs reviewed, the most common clinical trial design was superiority, used in 10 drugs (45.5%), followed by non-inferiority designs in 4 drugs (18.2%) and equivalence designs in 2 drugs (9.1%). Additionally, combined non-inferiority and equivalence designs were used in 2 drugs (9.1%). In one drug, the included pivotal trials employed a combined superiority and non-inferiority design (4.5%), and for another drug the main trial had not specified the design (4.5%). In terms of endpoints, the majority of drugs (13 out of 22; 59.1%) were assessed exclusively using surrogate endpoints. Clinical endpoints were used in 7 drugs (31.8%), while clinical scale endpoints were reported in 2 drugs (9.1%).

Of the 31 failed trials identified, 28 (90.3%) were RCTs. Regarding trial design, 17/31 (54.8%) were superiority studies, 7 (22.6%) non-inferiority studies, 6 (19.4%) equivalence studies, while 1 trial (3.2%) did not report its design. In terms of endpoints, 21 failed trials (67.7%) were based on surrogate measures, 8 (25.8%) on clinical outcomes, and 2 (6.5%) on clinical scales.

The explanations for approval despite failed endpoints varied widely and are detailed in Table 2. The most common reason for approval despite null results, for 9/22 (40.9%), was the approval based on the totality of evidence, secondary endpoints, or clinical judgment. Two (9.1%) were justified based on high unmet medical need or urgency. Another 2 (9.1%) were approved only after one of the initially proposed indications was withdrawn from the application, leading to approval for the remaining indications. Regulatory alignment with FDA standards justified approval for additional 2 (9.1%) therapeutics. One drug (4.5%), Truvelog Mix 30, was justified through post hoc analysis. Quviviq (4.5%) was partially approved with only the higher dose endorsed, and Hepcludex (4.5%) received conditional approval. For 4 therapeutics (18.2%) no justification was found in the EPAR.

Four of the 22 drugs (18.2%) had a total of six postmarketing requirement studies aimed at further assessing efficacy. In these cases, the requirements were linked to the regulatory pathway: one drug (Hepcludex) was authorised under conditional approval, while the other three were approved under exceptional circumstances.

## Discussion

### Main findings

Among the new drugs and biologics approved by the EMA between 2020 and 2023, there was substantial variability in the design and evidentiary strength of the supporting pivotal trials. While the majority of approvals were supported by strong trial designs, including 83% being RCTs, 59% of trials relied on surrogate endpoints. Moreover, nearly 10% of the approvals were based on pivotal trials that did not meet one of its primary endpoints. For a third of these drugs with missed primary endpoints, there were no pivotal trials demonstrating statistically significant efficacy. These findings show that even when designs are rigorous, the outcomes assessed may not always correspond directly to patient-relevant benefits.

### Handling of unmet primary endpoints

Despite the general expectation that pivotal trials meet their predefined primary endpoints, our findings show that this is not always the case. In some instances, the failed endpoint came from a supplemental trial among multiple studies. In such cases, the impact of a failed study may be mitigated by the presence of additional pivotal trials showing positive results. For example, the diabetes drug Rybelsus had two pivotal trials that failed to meet their primary endpoints, but six other trials in the same approval package demonstrated statistically significant benefits, as acknowledged by the EMA.

Approving a medicine despite a missed primary endpoint in a pivotal trial is exceptional, and the EMA’s assessment reports indicate that such approvals were granted only after careful consideration of additional evidence and context. Several patterns emerged in how EMA handled cases where primary endpoints were missed. First, the EMA occasionally accepted post hoc analyses or revised endpoints, provided strong justification existed. For example, biosimilar insulin Truvelog Mix 30 narrowly missed its non-inferiority margin for HbA1c reduction due to an outlier patient who mistakenly used expired study medication. Although the intention-to-treat analysis failed, regulators accepted a post hoc analysis excluding this patient, acknowledging the protocol violation and emphasizing that HbA1c was not critical for biosimilar approval. However, reliance on post hoc analyses, particularly by the EMA, is problematic, as such analyses are conducted after results are known and can also obscure potential risks. This pragmatic approach balanced scientific rigor with flexibility, ensuring benefit-risk was unaffected since the endpoint mainly supported similarity rather than novel benefit. Second, EMA sometimes narrowed the indication when efficacy was not proven across all intended populations. The COVID-19 prophylactic antibody Evusheld demonstrated strong efficacy pre-exposure but failed in asymptomatically infected patients (i.e. post-exposure setting). EMA resolved this by restricting approval solely to pre-exposure prophylaxis. Likewise, this happened for the influenza vaccine Fluad Tetra. The sponsor requested approval in two populations (pediatric and elderly). A Phase 3 trial in children failed to meet its endpoint and the requested pediatric indication was not granted by the EMA. Third, in scenarios of unmet medical need or public health significance, the agency made exceptions. As such, the COVID-19 vaccine Bimervax missed its primary immunogenicity endpoint against the reference virus strain. However, secondary endpoints showed strong responses against relevant newer variants, leading EMA to conclude the vaccine was approvable due to the epidemiological irrelevance of the original strain. Finally, the recent refusal of marketing authorisation for Nezglyal by the European Medicines Agency (EMA) reveals the inconsistencies that can arise in the interpretation of evidence across rare disease approvals. Nezglyal, which was developed for a neuromuscular condition, was evaluated in a single clinical trial that failed to show a clinically meaningful improvement on the 6-minute walk test, a commonly used endpoint in neuromuscular and metabolic disorders. Despite some signs of secondary benefit, the EMA concluded the evidence was insufficient and refused authorisation. This decision stands in contrast to other approvals where similarly limited or even negative trial results have been accepted. For example, Pombiliti, developed for Pompe disease, was approved despite not meeting its primary endpoint.^10^

### Trial Design and Endpoints in EMA authorisations

The reliability of drug approvals depends not only on whether the endpoints of pivotal trials are met, but on how they are designed and which outcomes they measure. Trial design and endpoint choices can accelerate development, but they also determine whether regulatory decisions translate into meaningful benefits for patients. In our sample, about 17% of pivotal trials were not RCTs, reflecting EMA’s occasional reliance on non-comparative evidence. Unlike the FDA, the EMA never required two RCTs for regulatory approval. However, the European regulator introduced in 2024 guidance on Single-Arm Trials (SATs)^11^. These may be accepted only in exceptional cases where RCTs are infeasible, though their lack of randomization increases bias. EMA therefore stresses careful design, robust endpoints, and the use of external comparators, while warning of their methodological limits.

Moreover, surrogate endpoints were used in well over half of the trials, and a majority of the approvals with failed endpoints relied on surrogate markers as primary endpoints. While such endpoints can expedite the approval process, their use has been widely criticized, as they often do not reliably predict meaningful clinical outcomes. This limitation is particularly evident in oncology, where surrogate measures frequently fail to translate into gains in overall survival or improvements in quality of life. A review of 68 oncology approvals (2009–2013) found only 35% with survival gains and 10% with quality-of-life benefits, despite most being based on surrogates.^12^ Nonetheless, this high number is in relation with prior research. For example, 90% of products approved via EMA’s special programs from 2011 to 2018 were based on surrogate primary endpoints.^13^

Non-inferiority trials also raised concerns, appearing in about one in five pivotal studies. Their validity depends on assumptions about historical controls and appropriately set margins; if poorly justified, they risk approving ineffective treatments.^14,15^

### Comparison of failed endpoints in a broader context

In general, our findings about EMA review outcomes mirror those reported in the literature and shed light on differences between EMA and FDA regulatory approaches. In the United States, the FDA has historically required “substantial evidence of effectiveness” from at least two adequate and well-controlled trials for approval, but this requirement can also be fulfilled by a single pivotal trial and confirmatory evidence, which may include observational studies, real-world evidence, natural history evidence and more. ^16,17^

Nonetheless, regarding missed primary endpoints, the FDA showed a similar percentage with 10% of drugs containing at least one primary failed endpoint.^4^ This trend appears even more pronounced in approvals related to high-risk therapeutic medical devices, where significant uncertainty persists regarding the robustness of efficacy data provided at approval.^18^

### Concept of negative trials

In regard of our results, it’s important to discuss the concept of a negative trial. The classification of a trial as positive or negative, follows the binary decision rule of Neyman and Pearson, in which the failure to reject the null hypothesis at a predefined significance level (usually p < 0.05) implies lack of efficacy.^19^ The latter framework has clear strengths such as that it is transparent, hypothesis-driven and offers a simple and consensual basis for decision-making. However, this binary model also has limitations. It may not fully capture the clinical relevance of borderline or consistent signals across multiple endpoints or populations. When a trial fails to meet its primary endpoint, it does not necessarily mean the intervention is ineffective. As our results show, regulatory agencies can occasionally approve drugs based on compelling secondary endpoints, subgroup analyses or alternative statistical approaches, especially when the totality of evidence suggests a clinically meaningful benefit. In some cases, these decisions appear justified and in line with the aim of ensuring timely patient access to beneficial therapies. Still, this flexibility opens the door to case-by-case exceptions that may blur the objectivity of the regulatory standard, especially when similar dossiers are treated inconsistently. Thus, while findings outside of primary endpoints may provide valuable insights, they must be interpreted with caution and in light of both methodological robustness and consistency across the evidence base.^20^

### Limitations

This study has several limitations that should be acknowledged. First, the analysis was restricted to EMA-approved drugs between 2020 and 2023, a period influenced by the COVID-19 pandemic and possibly atypical regulatory flexibility, limiting generalizability to other time frames or regulatory contexts and possibly raising the number of negative trials found in vaccine drugs. By focusing only on approved drugs and excluding withdrawn or refused applications, the study may introduce selection bias and potentially underestimate the frequency of trial failures or evidentiary weaknesses. Moreover, our assessments rely on information from EPARs, which, while comprehensive, are secondary sources and may lack full transparency or consistency. The lack of access to full study reports also limits the ability to independently assess trial quality or validate findings.

## Conclusion

The quality of clinical trial evidence supporting EMA approvals of novel therapeutic agents between 2020 and 2023 varied considerably. Notably, 1 in 10 of these approvals included at least one pivotal trial that failed to meet one of its primary endpoints. While these findings do not necessarily question the legitimacy of the approvals, they highlight the complexity of evidence evaluation in regulatory decision-making.

## Data Availability

Data will be shared upon publication of the article.

